# Monitoring sociodemographic inequality in COVID-19 vaccination coverage in England: a national linked data study

**DOI:** 10.1101/2021.10.07.21264681

**Authors:** Ted Dolby, Katie Finning, Allan Baker, Leigh Dowd, Kamlesh Khunti, Cameron Razieh, Thomas Yates, Vahé Nafilyan

**Affiliations:** Office for National Statistics, Newport, UK; Faculty of Public Health, Environment and Society, London School of Hygiene and Tropical Medicine, London, UK; Office for Health Improvement and Disparities, London, UK; Diabetes Research Centre, University of Leicester, Leicester General Hospital, Leicester, LE5 4PW, UK; National Institute for Health Research (NIHR) Leicester Biomedical Research Centre (BRC), Leicester General Hospital, Leicester, LE5 4PW, UK; University Hospitals of Leicester NHS Trust, Leicester, UK; Leicester Real World Evidence Unit, Diabetes Research Centre, University of Leicester, Leicester, UK

**Author notes:** These two authors contributed equally to this paper (Ted Dolby, Katie Finning). Corresponding author: Katie Finning.

## Abstract

**Background:** The UK began an ambitious COVID-19 vaccination programme on 8^th^ December 2020. This study describes variation in vaccination coverage by sociodemographic characteristics between December 2020 and August 2021.

**Methods:** Using population-level administrative records linked to the 2011 Census, we estimated monthly first dose vaccination rates by age group and sociodemographic characteristics amongst adults aged 18 years or over in England. We also present a tool to display the results interactively.

**Findings:** Our study population included 35,223,466 adults. A lower percentage of males than females were vaccinated in the young and middle age groups (18-59 years) but not in the older age groups. Vaccination rates were highest among individuals of White British and Indian ethnic backgrounds and lowest among Black Africans (aged ≥80 years) and Black Caribbeans (18-79 years). Differences by ethnic group emerged as soon as vaccination roll-out commenced and widened over time. Vaccination rates were also lower among individuals who identified as Muslim, lived in more deprived areas, reported having a disability, did not speak English as their main language, lived in rented housing, belonged to a lower socio-economic group, and had fewer qualifications.

**Interpretation:** We found inequalities in COVID-19 vaccination rates by sex, ethnicity, religion, area deprivation, disability status, English language proficiency, socio-economic position, and educational attainment, but some of these differences varied by age group. Research is urgently needed to understand why these inequalities exist and how they can be addressed.

**Research in context:** *Evidence before this study:* We searched PubMed for publications on sociodemographic inequalities in COVID-19 vaccination coverage. Several studies have reported differences in coverage by characteristics such as ethnicity and religion, however these have focused on older adults and the clinically vulnerable who were initially prioritized for vaccination. There is little evidence on sociodemographic inequalities in vaccination coverage among younger adults and evidence is also lacking on coverage by a wider range of characteristics such as sex, disability status, English language proficiency, socio-economic position, and educational attainment.

*Added value of this study:* This study provides the first evidence for sociodemographic inequalities in COVID-19 vaccination coverage among the entire adult population in England, using population-level administrative records linked to the 2011 Census. By disaggregating the data by age group, we were able to show that disparities in coverage by some sociodemographic characteristics differed by age group. For example, a lower proportion of males than females were vaccinated in the young and middle age groups (18-59 years) but not in the older age groups, and vaccination rates were lowest among Black Africans in those aged ≥80 years but lowest among Black Caribbeans for all other age groups. Vaccination rates were also lower among individuals who identified as Muslim, lived in more deprived areas, reported having a disability, did not speak English as their main language, lived in rented housing, belonged to a lower socio-economic group, and had fewer qualifications.

*Implications of all the available evidence:* Many of the groups with the lowest rates of COVID-19 vaccination are also the groups that have been disproportionately affected by the pandemic, including severe illness and mortality. Research is urgently needed to understand why these disparities exist and how they can be addressed, for example through public health or community engagement programmes. Since the relationships between sociodemographic characteristics and vaccination coverage may differ by age group, it is important for future research to disaggregate by age group when examining these inequalities.

## Introduction

The UK began a universal vaccination programme to combat the COVID-19 pandemic on 8th December 2020; by 27 September 2021, 89.6% of the UK adult population had received their first dose [1].

Previous research demonstrates that rates of vaccination for a variety of diseases are lower amongst certain ethnic groups and in areas of higher deprivation [2, 3, 4, 5]. Evidence also suggests that rates of COVID-19 vaccination differ by sociodemographic factors, religion, and certain underlying health conditions [6, 7]. However, the evidence for COVID-19 vaccination rates by characteristics has so far focused on older adults and the clinically vulnerable, who were initially prioritised for vaccination. Less is known about COVID-19 vaccination rates in younger adults, or about differences in the impact of sociodemographic factors across different age groups. Understanding which sociodemographic, economic, and cultural factors are associated with lower vaccination rates across the whole adult population has major implications for the design of policies that help to maximise coverage of the vaccination campaign. Achieving a high rate of vaccination in the population and not just in those at the highest risk is critical to slow infection, reduce hospital admissions, and help healthcare systems and countries recover from the pandemic [8].

This study investigates inequality in vaccination rates by age group and sociodemographic characteristics amongst adults aged 18 years or over in England, using population-level administrative records linked to the 2011 Census. This dataset enables examination of a wide range of sociodemographic characteristics lacking in previously published studies including age group, sex, ethnic group, religious affiliation, area deprivation, disability status, English language proficiency, National Statistics Socio-Economic Classification, and educational attainment.

## Methods

### Data

We linked vaccination data from the NHS England and NHS Improvement’s National Immunisation Management System (NIMS) to the Office for National Statistics (ONS) Public Health Data Asset (PHDA) based on NHS number, which is a unique identifier. The ONS PHDA is a linked dataset that includes the 2011 Census, mortality records, and the General Practice Extraction Service (GPES) data for pandemic planning and research. To obtain NHS numbers for the 2011 Census, we linked the 2011 Census to the 2011-2013 NHS Patient Registers using deterministic and probabilistic matching, with an overall linkage rate of 94.6%. All subsequent linkages were performed based on NHS numbers.

The study population consisted of adults aged 18 years or over, alive on 8 December 2020, who were resident in England, registered with a general practitioner, and enumerated at the 2011 Census. Of 38,066,935 adults aged 18 years or over who received a first dose of a COVID-19 vaccine in NIMS, 30,505,356 (80.1%) were linked to the ONS PHDA.

### Outcome

The main outcome was having received at least a first dose of a COVID-19 vaccine as recorded in the NIMS data available on 15 September 2021. To calculate monthly rate, vaccination status was assessed on the last day of the month.

### Exposures

This dataset contains comprehensive sociodemographic information from the 2011 Census and geographical information from GPES. Demographic and socio-economic characteristics including age, sex, ethnic group, religious affiliation, socio-economic status, and self-reported disability status were based on the 2011 Census. We used a 10-category ethnic group classification (White British, Bangladeshi, Black African, Black Caribbean, Chinese, Indian, Mixed, Other, Pakistani, White Other). Geographical factors (region, urban/rural status) and area deprivation (Index of Multiple deprivation, IMD [9]) were derived from the 2019 GPES. We also considered English language proficiency, educational attainment, the National Statistics Socio-Economic Classification (NS-SEC) and household tenure, all of which were drawn from the 2011 Census. A list of all variables included in this analysis and their source is provided in Table 1.

**Table 1.** Variables used in the analyses

| Variable | Coding | Source |
| --- | --- | --- |
| Vaccinated | Received a first dose of a COVID-19 vaccine by 31<sup>st</sup> August 2021 | NIMS |
| Age | Third-order polynomial | 2011 Census |
| Sex | Female, Male | 2011 Census |
| Ethnicity | White British, Bangladeshi, Black African, Black Caribbean, Chinese, Indian, Mixed, Other, Pakistani, White other | 2011 Census |
| Religious affiliation | Christian, Buddhist, Hindu, Jewish, Muslim, Sikh, Other Religion, No religion, Religion Not Stated | 2011 Census |
| English language proficiency | Main language, Not main language | 2011 Census |
| Region | Dummy variables representing region of residence | GPES |
| Rural urban classification | Urban, rural | GPES |
| Index of Multiple Deprivation (IMD) | Dummy variables representing quintiles of deprivation | GPES |
| Household tenure | Own, social rented, private rented, other | 2011 Census |
| Level of highest qualification | Level 4+ (Degree or equivalent), Level 3 (A-level or equivalent), Apprenticeship, Level 2 (GCSE or equivalent), Level 1, other, No qualification | 2011 Census |
| National Statistics Socio-economic classification (NS-SEC) | Higher managerial, administrative and professional occupations, Lower managerial, administrative and professional occupations, Intermediate occupations, Small employers and own account workers, Lower supervisory and technical occupations, Semi-routine occupations, Routine occupations, Never worked and long-term unemployed, Not classified | 2011 Census |
| Disability | Non-disabled, disabled (limited a little), disabled (limited a lot) | 2011 Census |

GPES - General Practice Extraction Service; NIMS - National Immunisation Management System

### Statistical analyses

We estimated monthly first dose vaccination rates by age group and a range of demographic and socio-economic characteristics. For every month between December 2020 and August 2021 and for each age group, we estimated the rate of people who had received at least a first dose of a vaccination against COVID-19 by the end of the month, restricting the sample to people alive at the end of the specified month. Plots were produced to provide a visual summary of vaccination coverage over time by demographic and socio-economic characteristics. Analyses were conducted using R version 3.5.

### Data sharing statement

The monthly vaccination rates by age group and the full list of demographic and socio-economic characteristics used in this paper are displayed on the COVID-19 Health Inequalities Monitoring for England (CHIME) tool. The CHIME tool displays the number and proportion of people who have not yet received their first dose of COVID-19 vaccination. For the analysis presented here we derived first dose vaccination rates by subtracting the number who had not received their first dose from the denominator.

### Role of the funding source

The funding source played no part in the interpretation of the results.

## Results

### Characteristics of study population

Our study population included 35,223,466 adults aged 18 years or over who lived in England. Of these, 52.4% were female, 82.4% identified as White British, 60.5% identified as Christian, and 14.5% reported having a disability and being limited a little (8.8%) or a lot (5.7%) in their daily activities. Table 2 provides detailed characteristics of the sample.

**Table 2.** Characteristics of the study population

| Characteristic | Level | Number (%) |
| --- | --- | --- |
| Sex | Female | 18,455,496 (52.4) |
|  | Male | 16,767,970 (47.6) |
| Age (years) | 18-29 | 6,024,663 (17.1) |
|  | 30-39 | 5,196,626 (14.8) |
|  | 40-49 | 5,561,817 (15.8) |
|  | 50-59 | 6,430,167 (18.3) |
|  | 60-69 | 5,234,144 (14.9) |
|  | 70-79 | 4,263,033 (12.1) |
|  | 80+ | 2,513,016 (7.1) |
| Ethnicity | Bangladeshi | 270,035 (0.8) |
|  | Black African | 538,586 (1.5) |
|  | Black Caribbean | 378,063 (1.1) |
|  | Chinese | 187,646 (0.5) |
|  | Indian | 946,289 (2.7) |
|  | Mixed | 584,822 (1.7) |
|  | Other | 858,255 (2.4) |
|  | Pakistani | 708,387 (2.0) |
|  | White British | 29,024,719 (82.4) |
|  | White other | 1,726,664 (4.9) |
| Religion | Buddhist | 146,584 (0.4) |
|  | Christian | 21,309,133 (60.5) |
|  | Hindu | 537,242 (1.5) |
|  | Jewish | 159,557 (0.5) |
|  | Muslim | 1,598,295 (4.5) |
|  | Sikh | 292,093 (0.8) |
|  | Other religion | 162,467 (0.5) |
|  | No religion | 8,808,882 (25.0) |
|  | Religion not stated | 2,209,213 (6.3) |
| Disability Status | Not limited | 30,095,777 (85.4) |
|  | Limited a little | 31,101,61 (8.8) |
|  | Limited a lot | 2,017,528 (5.7) |
| Index of Multiple Deprivation (IMD) Quintile | 1 (Most deprived) | 6,487,968 (18.4) |
|  | 2 | 6,875,573 (19.5) |
|  | 3 | 7,186,492 (20.4) |
|  | 4 | 7,325,184 (20.8) |
|  | 5 (Least deprived) | 7,348,249 (20.9) |
| Household Tenure | Owned | 24,190,067 (68.7) |
|  | Private rented | 4,991,430 (14.2) |
|  | Social rented | 5,183,538 (14.7) |
|  | Other | 556,343 (1.6) |
|  | Not classified | 302,088 (0.9) |
| English Language Proficiency | Main language | 32,762,210 (93.0) |
|  | Not main language | 2,461,256 (7.0) |
| Educational Attainment | Level 4+ | 8,866,351 (30.4) |
|  | Level 3 | 3,608,197 (12.4) |
|  | Apprenticeship | 1,061,770 (3.6) |
|  | Level 2 | 4,326,004 (14.8) |
|  | Level 1 | 4,065,820 (13.9) |
|  | Other | 1,593,068 (5.5) |
|  | No qualification | 5,677,593 (19.4) |
| National Statistics Socio-Economic Classification (NS-SEC) | 1 Higher managerial, administrative and professional occupations | 4,957,652 (14.1) |
|  | 2 Lower managerial, administrative and professional occupations | 8,210,886 (23.3) |
|  | 3 Intermediate occupations | 3,687,464 (10.5) |
|  | 4 Small employers and own account workers | 4,651,919 (13.2) |
|  | 5 Lower supervisory and technical occupations | 3,086,562 (8.8) |
|  | 6 Semi-routine occupations | 4,454,670 (12.6) |
|  | 7 Routine occupations | 4,150,251 (11.8) |
|  | 8 Never worked and long-term unemployed | 1,209,877 (3.4) |
|  | Not classified | 814,185 (2.3) |

Note: Adults aged 18 years and over living in England, alive on 8th December 2020.

### Inequality in vaccination coverage

Figures 1 to 3 show the proportion of people vaccinated over time by age group and: sex (Figure 1), ethnic group (Figure 2), and religion (Figure 3). All underlying data can be interactively accessed through the COVID-19 Health Inequalities Monitoring for England (CHIME) tool. When examining all age groups combined, vaccination coverage was higher among females compared to males (Figure 1). When stratified by age group this difference between sexes was pronounced among younger adults, particularly the 18-29, 30-39, 40-49 and, to a lesser extent, 50-59 year old age groups. Amongst those aged 60-69, 70-79, and 80 years and over, vaccination rates were approximately equivalent for males and females (Figure 1). The sex difference observed in the younger age groups was widest in the earlier months of vaccination roll-out and has since narrowed but has not been eliminated (Figure 1). As at the end of August 2021, 24% of females aged 18-29 years and 21% of females aged 30-39 years had not received their first vaccination, compared with 29% and 25% of males in the same age groups, respectively.

**Figure 1.**
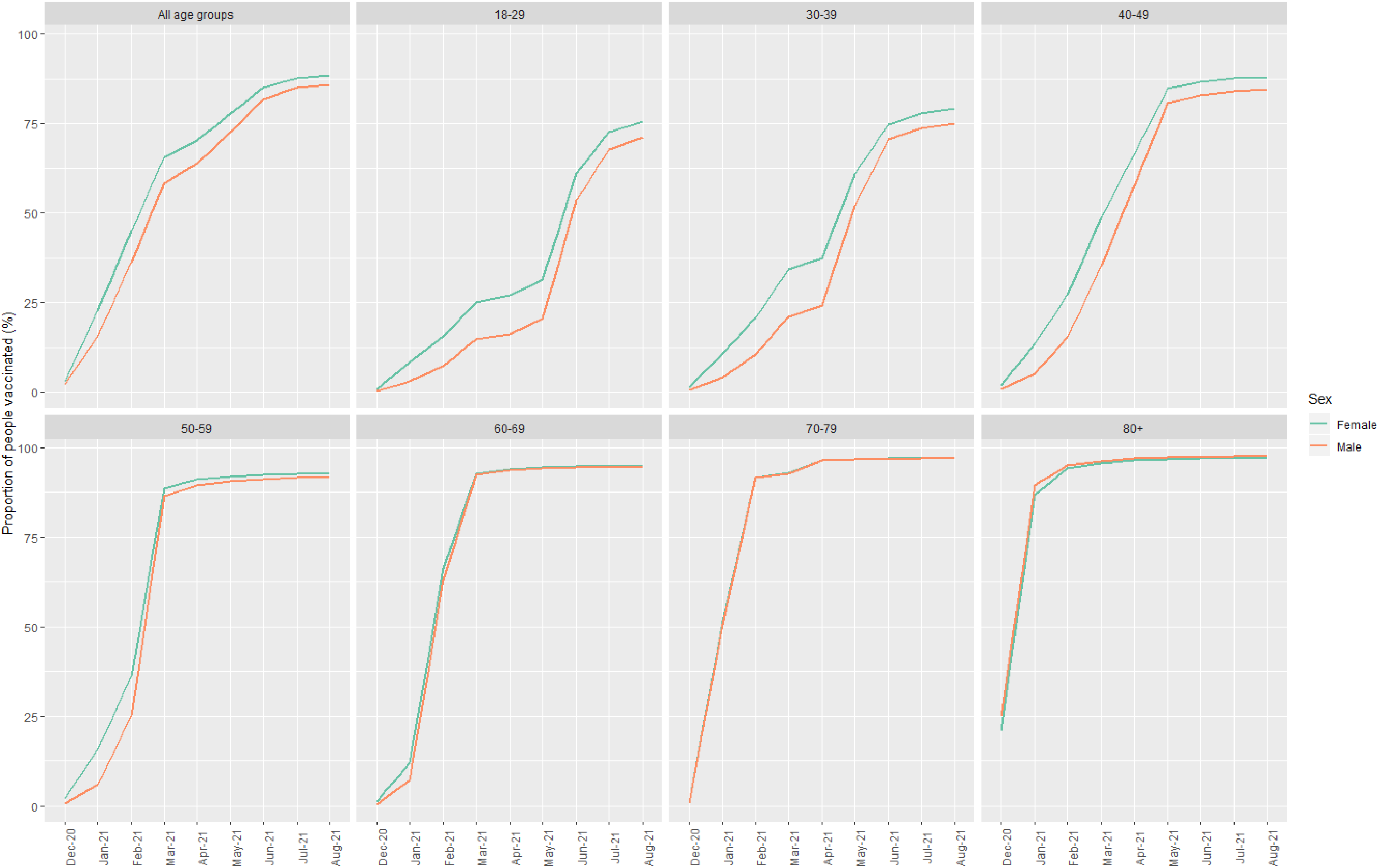
Proportion of people who received at least one dose of COVID-19 vaccination over time, by sex and age group

**Figure 2.**
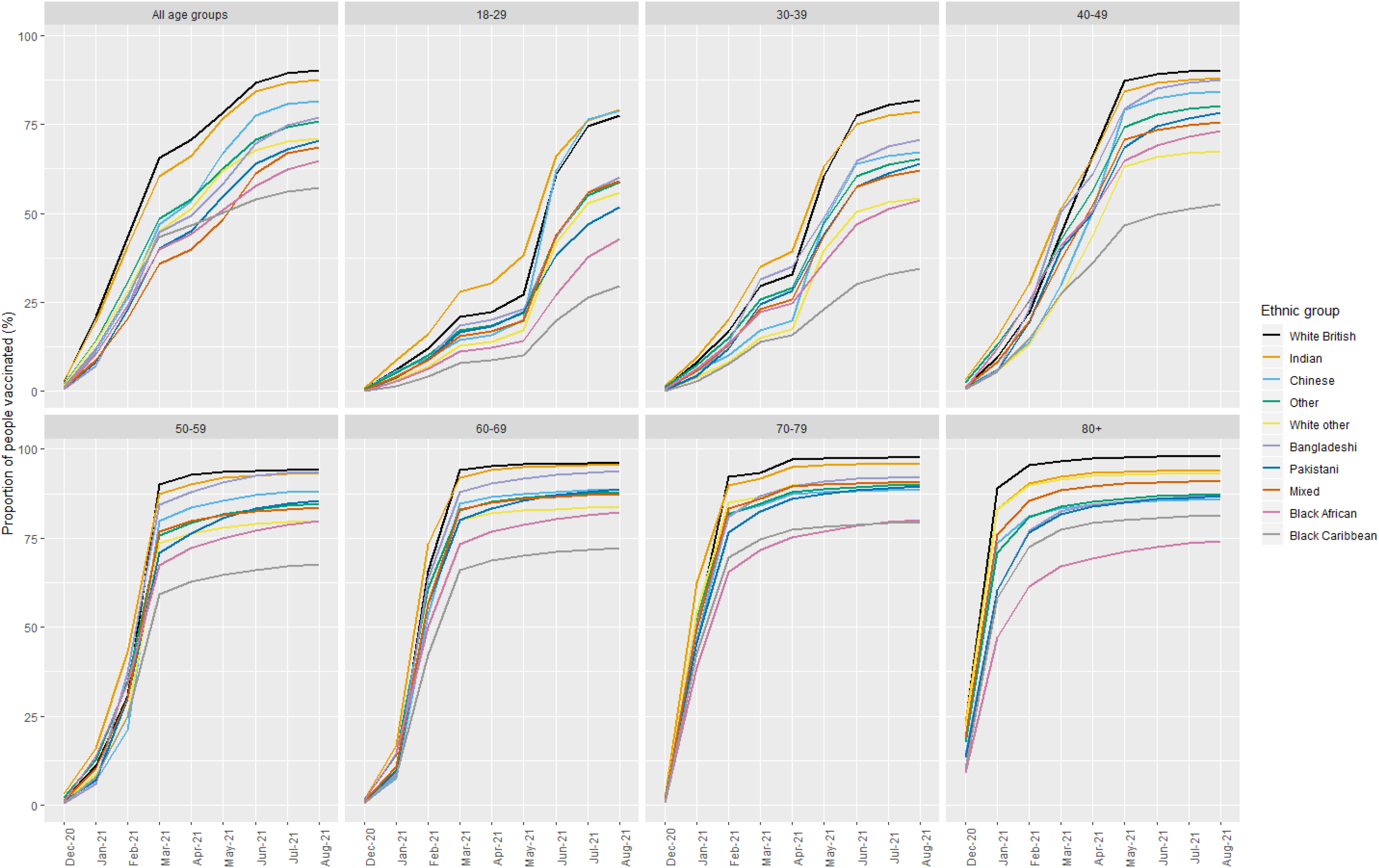
Proportion of people who received at least one dose of COVID-19 vaccination over time, by ethnic group and age group

**Figure 3.**
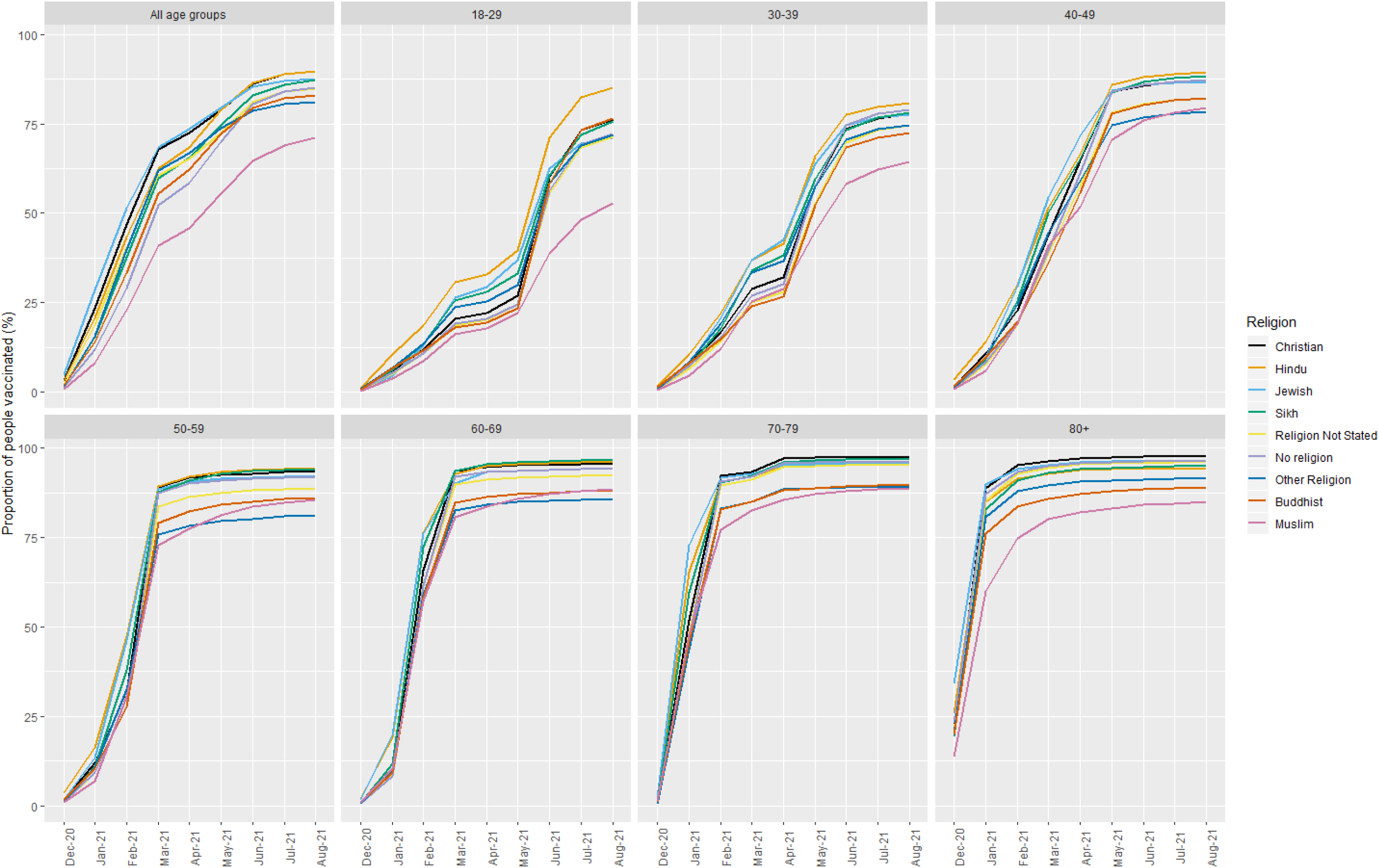
Proportion of people who received at least one dose of COVID-19 vaccination over time, by religion and age group

In all age groups combined, vaccination rates were highest among White British and Indian ethnic groups, and lowest among Black Caribbean, Black African, Mixed, and Pakistani ethnic groups (Figure 2). However, these ethnic differences varied according to age group. For example, among those aged 18-29 years, vaccination rates were highest for those identifying as Indian followed by Chinese and White British; whereas in the 80 years and over age group, coverage was greatest for those identifying as White British followed by Indian and White Other. In the 80 years and over age group coverage was lowest for individuals of Black African ethnic background, but among all other age groups coverage was lowest for individuals of Black Caribbean ethnic background. Differences in vaccination coverage by ethnic group emerged as soon as vaccination roll-out began within each age group and tended to widen over time (Figure 2). As at the end of August 2021, only 57% of adults in England identifying as Black Caribbean had received at least one dose of COVID-19 vaccination, compared with 90% of adults identifying as White British and 88% of those identifying as Indian.

Vaccination rates also varied across religious groups (Figure 3). When all age groups were combined, individuals identifying as Muslim had markedly lower vaccination rates than other religious groups, while those belonging to Hindu and Christian religious groups had the highest vaccination coverage. When stratified by age group, adults identifying as Christian had the highest rates of vaccination among those aged 70-79 and 80 years and over, but in the younger age groups coverage among Christians was comparatively lower and was greatest among those identifying as Hindu (Figure 3). As at the end of August 2021, only 71% of adults identifying as Muslim had received at least one dose of COVID-19 vaccination, compared with 90% of adults identifying as Hindu or Christian.

Vaccination coverage also differed by area deprivation and disability status (see Figures 4 and 5). In the early months of vaccination roll-out for each age group, uptake rates followed similar trajectories for the different IMD groups, but differences emerged in later months, with coverage being lowest among those living in the most deprived areas (IMD quintile 1) and highest among those in the least deprived areas (IMD quintile 5) (Figure 4). These differences by IMD quintile were greater among the younger age groups. As at the end of August 2021, 40% of individuals aged 18-29 years living in the most deprived areas had not been vaccinated, compared with 17% of adults of the same age living in the least deprived areas. For individuals aged 80 years and over these percentages were 5% and 2%, respectively.

**Figure 4.**
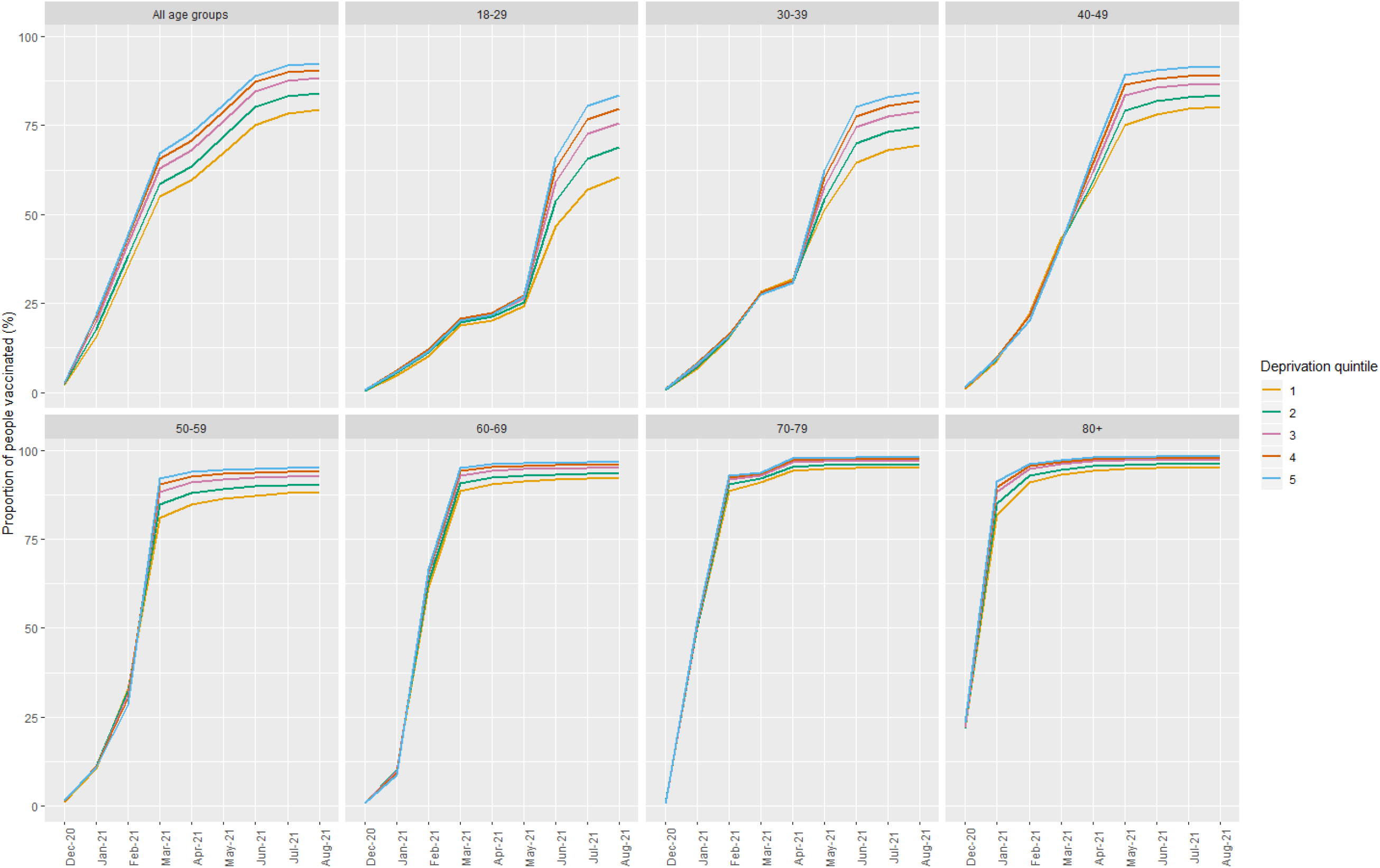
Proportion of people who received at least one dose of COVID-19 vaccination over time, by Index of Multiple Deprivation (IMD) quintile and age group. Note: IMD quintile 1 indicates those living in the most deprived areas and quintile 5 indicates those living in the least deprived areas.

**Figure 5.**
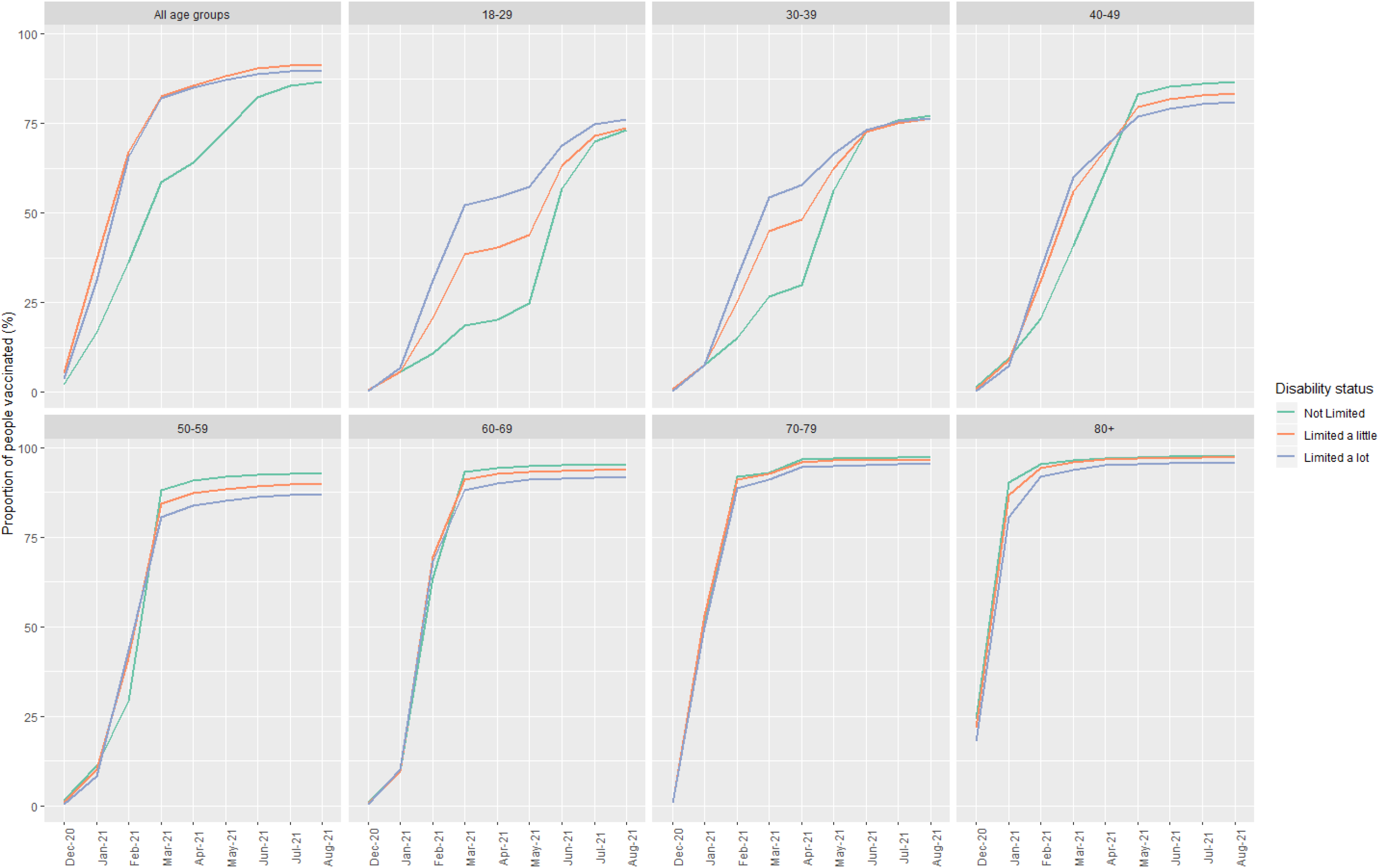
Proportion of people who received at least one dose of COVID-19 vaccination over time, by disability status and age group

Across most age groups, vaccination rates were initially higher for individuals with a disability compared to those with no disability (Figure 5). However, vaccination rates among those with no disabilities increased in later months, and once coverage plateaued within each age group the overall proportion of people vaccinated was marginally lower among those with disabilities (Figure 5).

Vaccination rates also differed by English language proficiency, household tenure, NS-SEC, and educational attainment (see Supplementary Material Figures S1 to S4). In all age groups a greater proportion of individuals who speak English as their main language had been vaccinated compared to those who did not speak English as their main language (Figure S1). Among all age groups vaccination coverage was highest for adults who own their own home (Figure S2). Coverage was lowest in those living in social rented housing among the 18-29 and 30-39 year old age groups and lowest in those living in private rented housing among the 50 years and over age groups.

Vaccination rates were highest among adults in NS-SEC group 1 (higher managerial, administrative and professional occupations) and lowest among those in NS-SEC group 8 (never worked and long-term unemployed) (Figure S3). As at the end of August 2021, 92% of adults in NS-SEC group 1 had received at least their first dose of COVID-19 vaccination compared with 70% of adults in NS-SEC group 8. Finally, there were differences in vaccination coverage by educational attainment, although this varied according to age group (Figure S4). In the youngest age groups individuals in attainment categories level 3 (A-level or equivalent) and level 4+ (degree or equivalent) had the highest vaccination rates, whereas in the 50-59 and older age groups vaccination rates were highest among those with an apprenticeship. Among all age groups vaccination coverage was lowest in those with “other” qualifications, and as at the end of August 2021, 83% of adults in England whose highest qualification was “other” had been vaccinated compared with 93% of adults with an apprenticeship and 91% of those with a degree or equivalent.

## Discussion

### Main findings of this study

Using whole population level linked data in England, our analysis demonstrates that first dose vaccination rates in adults aged 18 years and over differed by sex, ethnic group, religious affiliation, area deprivation, disability status, English language proficiency, household tenure, NS-SEC, and educational attainment. In addition, some of these differences varied by age group. A lower percentage of males than females were vaccinated in the young and middle age groups (18-59 years) but not in the older age groups. Vaccination rates were highest among individuals of White British and Indian ethnic backgrounds, in addition to younger adults identifying as Chinese. Among individuals aged 80 years and over, coverage was lowest among Black Africans, but in all other age groups coverage was lowest for Black Caribbeans. Individuals identifying as Muslim had the lowest vaccination rates across all age groups. In those aged 70 years and over, those identifying as Christian were the most vaccinated, but in younger age groups uptake was comparatively lower among Christians and highest in those identifying as Hindu. Across all age groups vaccination coverage was lower among adults who lived in more deprived areas, reported having a disability, did not speak English as their main language, lived in rented accommodation, belonged to a lower National Statistics Socio-Economic Classification (NS-SEC) group, and had fewer qualifications.

### What is already known on this topic

Lower rates of vaccination among ethnic minority groups, particularly Black ethnic groups, have been reported for a variety of diseases including COVID-19, although studies on the latter have so far focused on older adults [2, 3, 4, 6, 7]. A study of seasonal influenza vaccination in England found that uptake was highest among Asian adults aged 18-64 [4]. While different ethnic categories were used in the current study, we did find high coverage among Indian and younger Chinese adults. Religion was recognised as a potential factor in vaccination behaviour prior to COVID-19 [10] and two recent analyses of COVID-19 vaccination among older adults in England found that, after adjusting for geographical and sociodemographic factors, coverage was lowest amongst individuals identifying as Muslim, Buddhist, or Other Religion [6, 11]. Greater area deprivation has been associated with lower rates of vaccination in general, and against COVID-19 specifically, although again evidence for the latter has been limited to older adults [2, 3, 4, 5, 6, 7]. Few studies have explored COVID-19 vaccination coverage by sex or disability status. One study of adults aged 80 years and over reported no difference in coverage by sex, and slightly higher coverage among those with physical comorbidities [7]. Likewise, a recent study reported slightly higher rates of vaccine coverage among adults aged 70 years and over with a disability compared to those without [6].

### What this study adds

We provided novel evidence for COVID-19 vaccination rates by sociodemographic characteristics covering the entire adult population in England. Unlike previous studies, we disaggregated coverage by age group in addition to other sociodemographic characteristics. Our data show that disparities in vaccination coverage by sociodemographic characteristics differ according to age group, which highlights the importance of separating age groups when examining vaccination coverage.

While previous studies have shown lower vaccination rates among Black ethnic groups in general, we demonstrate that the lowest coverage was for Black Africans in the 80 years and over age group, and Black Caribbeans in all other age groups. We also found, for the first time, high rates of vaccination among young adults belonging to Indian and Chinese ethnic groups. Our results add to mounting evidence that vaccination coverage is particularly low among Muslim individuals.

Like previous research, we found no evidence for sex differences in vaccination rates among older adults. However, by disaggregating our data by age group we were able to demonstrate for the first time that there were sex differences between the ages of 18 and 59, with greater coverage among females compared to males in this age group. This sex difference was greatest in the initial months of roll-out of the vaccination programme, which may be due to the greater proportion of women in health and social care roles who were initially prioritised for vaccination. However, this sex difference has not been eliminated over time. Our findings are consistent with previous studies showing that vaccination coverage is lower among people with disabilities and in those living in more deprived areas, however we additionally found that the differences in coverage by area deprivation appeared to be greater among younger adults.

### Strengths and limitations

A major strength of this study is the use of nationwide linked population-level data from clinical records and the 2011 Census. Unlike studies based solely on electronic health records, we were able to examine a wide range of sociodemographic characteristics; and unlike surveys, we were able to precisely estimate vaccination rates for small groups. Unlike previous research that has focused on initial months of the vaccination programme in England and is therefore limited to certain groups such as older adults and the clinically vulnerable, our data spans the entire vaccination programme between December 2020 and August 2021 and is therefore more representative of the whole adult population. This also enabled us to examine vaccination rates by age group in addition to other sociodemographic characteristics.

Another strength of this study is the publication of up-to-date vaccination rates broken down by sociodemographic characteristics on the COVID-19 Health Inequalities Monitoring for England (CHIME) tool. The tool provides the opportunity for users to monitor inequalities in vaccination rates over time, as it will be updated with new data every month. This paper presents the data used in this new tool, which is a key part of the surveillance system designed to help the COVID-19 policy response.

A limitation of this study is that most of the demographic and socio-economic characteristics were derived from the 2011 Census and are therefore 10 years old. We focused primarily on characteristics that are unlikely to change over time, such as ethnicity and religion, but for characteristics that are more likely to change over time, such as disability status, the time difference may introduce some bias. However, we would expect this to dilute any differences observed rather than over-inflate them, because we will be missing new disabilities. Area deprivation was derived from GPES and measured in 2019 and is therefore not subject to the same bias. An additional limitation is that, because the Public Health Data Asset was based on the 2011 Census, it excluded people who were living in England in 2011 but who did not take part in the Census, as well as respondents who could not be linked to the 2011-2013 NHS patient register and recent migrants. As a result, we excluded 19.9% of vaccinated people who could not be linked to the PHDA, therefore our population may not be fully representative of the population living in England. Finally, as our aim was to present differences in vaccination rates in order to identify areas of greatest public health priority, our analyses were descriptive in nature and stratified by, but did not adjust for, likely confounders. A previous study in older adults showed that adjusting for covariates reduced, but did not eliminate, differences in COVID-19 vaccination coverage by ethnic group, religious affiliation, and area deprivation [6].

## Conclusion

There are differences in COVID-19 vaccination rates over time by sex, ethnic group, religious affiliation, area deprivation, disability status, English language proficiency, socio-economic position, and educational attainment, but some of these differences vary by age group, highlighting the importance of disaggregating age groups when examining vaccination coverage. Many of the groups with lowest vaccination coverage are the ones that have been disproportionately affected by the pandemic, including severe illness and mortality, and research is urgently needed to understand why these disparities exist and how they can be addressed, for example through public health or community engagement programmes.

## Data Availability

https://analytics.phe.gov.uk/apps/chime/

## Acknowledgements

KK is supported by the National Institute for Health Research (NIHR) Applied Research Collaboration East Midlands (ARC EM) and the NIHR Leicester Biomedical Research Centre (BRC).

## Ethical Approval Statement

Ethical approval was obtained from the National Statistician’s Data Ethics Advisory Committee (NSDEC(20)12).

## Authorship statement

Study conceptualisation was led by VN, TD and KF. VN, TD, KF, AB and LD contributed to the development of the research question, study design, with development of statistical aspects led by TD and VN. TD and VN were involved in data specification, curation and collection. TD, KF and VN conducted and checked the statistical analyses. AB and LD developed the tool to visualise the results. All authors contributed to the interpretation of the results. KF and VN wrote the first draft of the paper. TD, TY, CR, KK, AB contributed to the critical revision of the manuscript for important intellectual content. All authors approved the final version of the manuscript. VN had full access to all data in the study and takes responsibility of the integrity of the data and the accuracy of the data analysis. The lead authors (TD and KF) affirm that the manuscript is an honest, accurate, and transparent account of the study being reported; that no important aspects of the study have been omitted; and that any discrepancies from the study as planned have been explained.

## Declaration of interests

KK is Chair of the Ethnicity Subgroup of the UK Scientific Advisory Group for Emergencies (SAGE) and Member of SAGE.

## Funding

This research was funded by ONS and PHE. TY KK and CR are supported by a grant from the UKRI (MRC)- DHSC (NIHR) COVID-19 Rapid Response Rolling Call (MR/V020536/1) and from HDR-UK (HDRUK2020.138).

## Notes

### Author Declarations

This study involves only openly available human data, which can be obtained from the COVID-19 Health Inequalities Monitoring for England (CHIME) tool https://analytics.phe.gov.uk/apps/chime/

