## Supplementary material for "Monitoring sociodemographic inequality in COVID-19 vaccination coverage in England: a national linked data study"

Figure S1. Proportion of people who received at least one dose of COVID-19 vaccination over time, by English language proficiency and age group

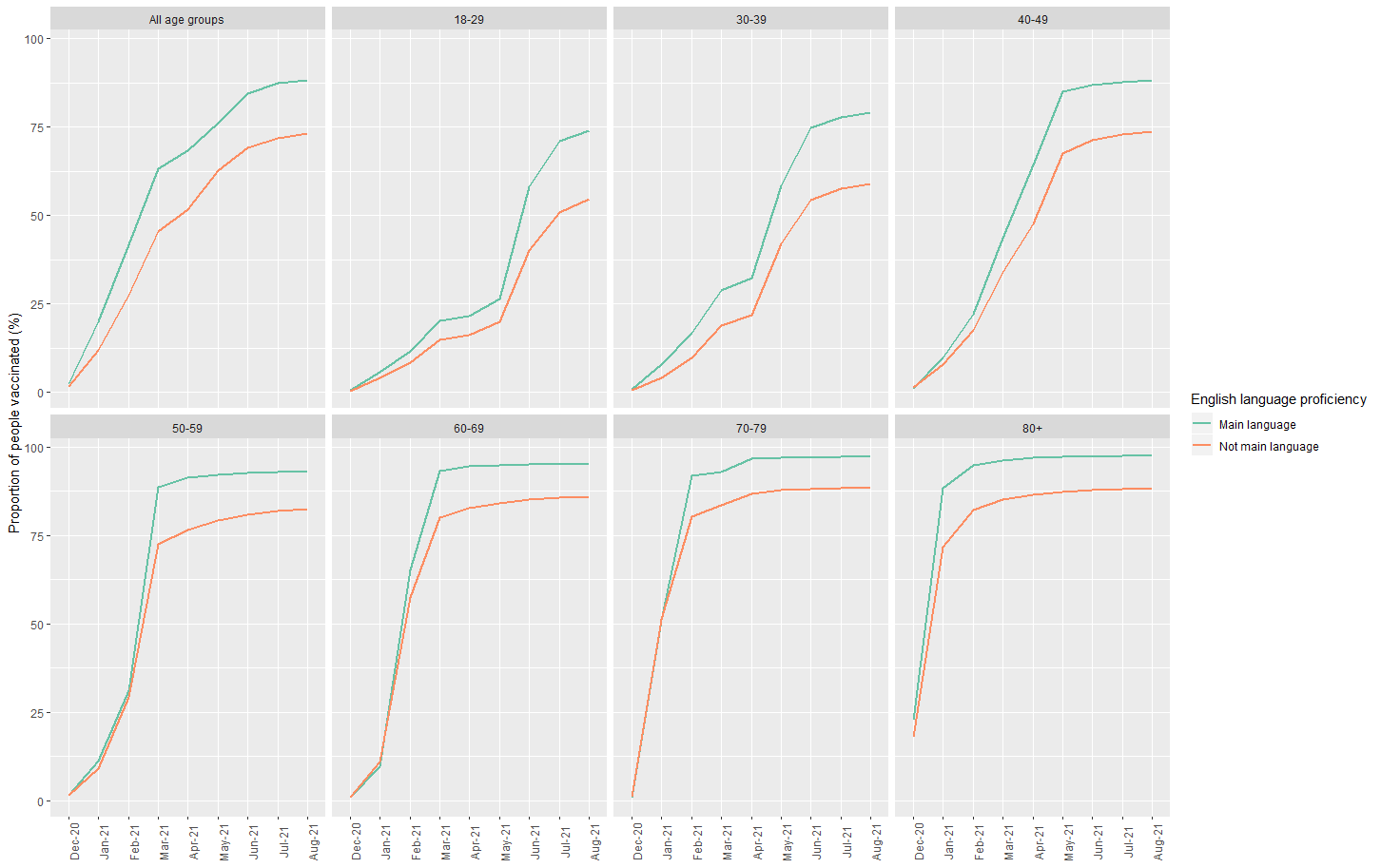

Figure S2. Proportion of people who received at least one dose of COVID-19 vaccination over time, by household tenure and age group

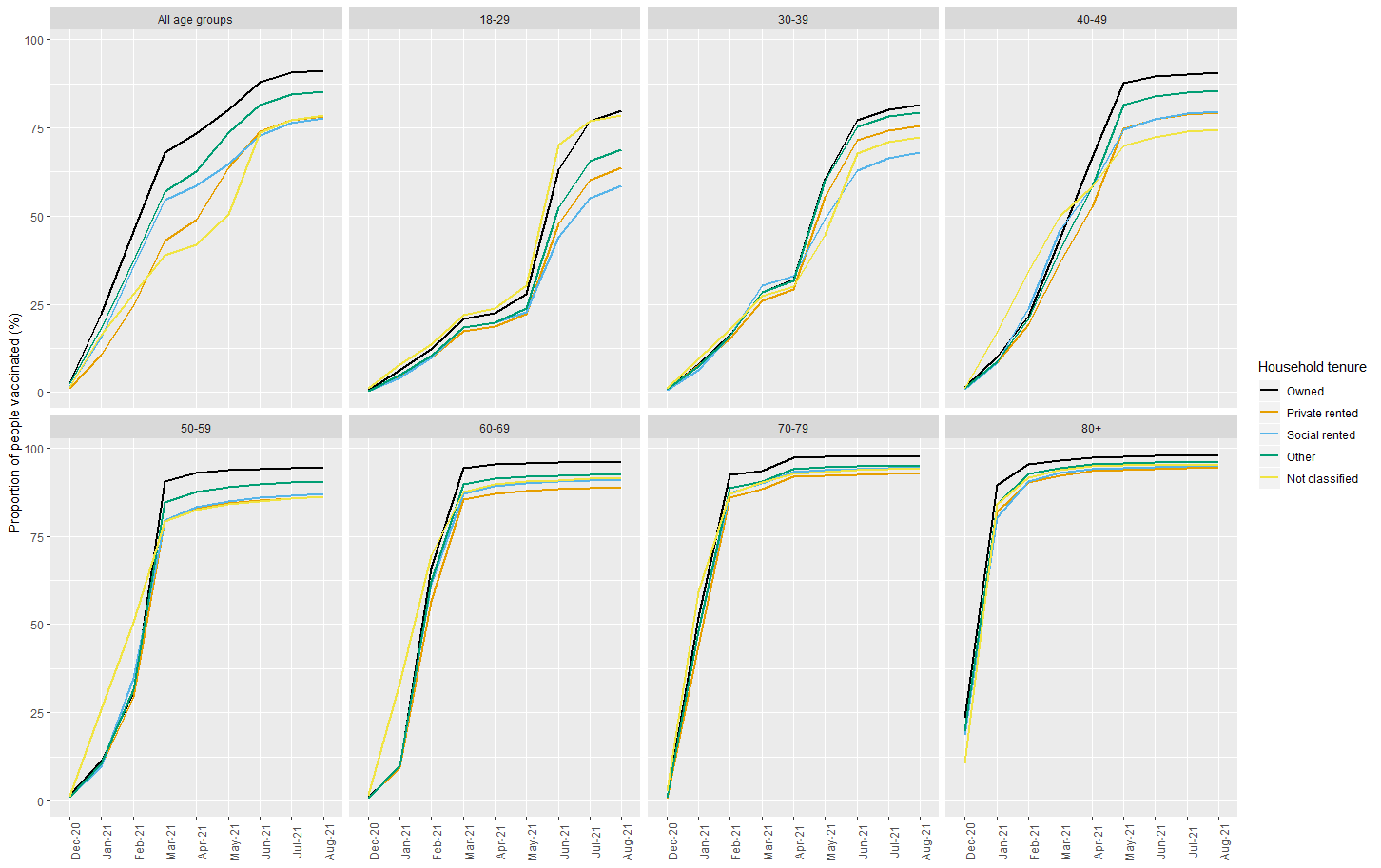

Figure S3. Proportion of people who received at least one dose of COVID-19 vaccination over time, by National Statistics Socio-Economic Classification (NS-SEC) and age group

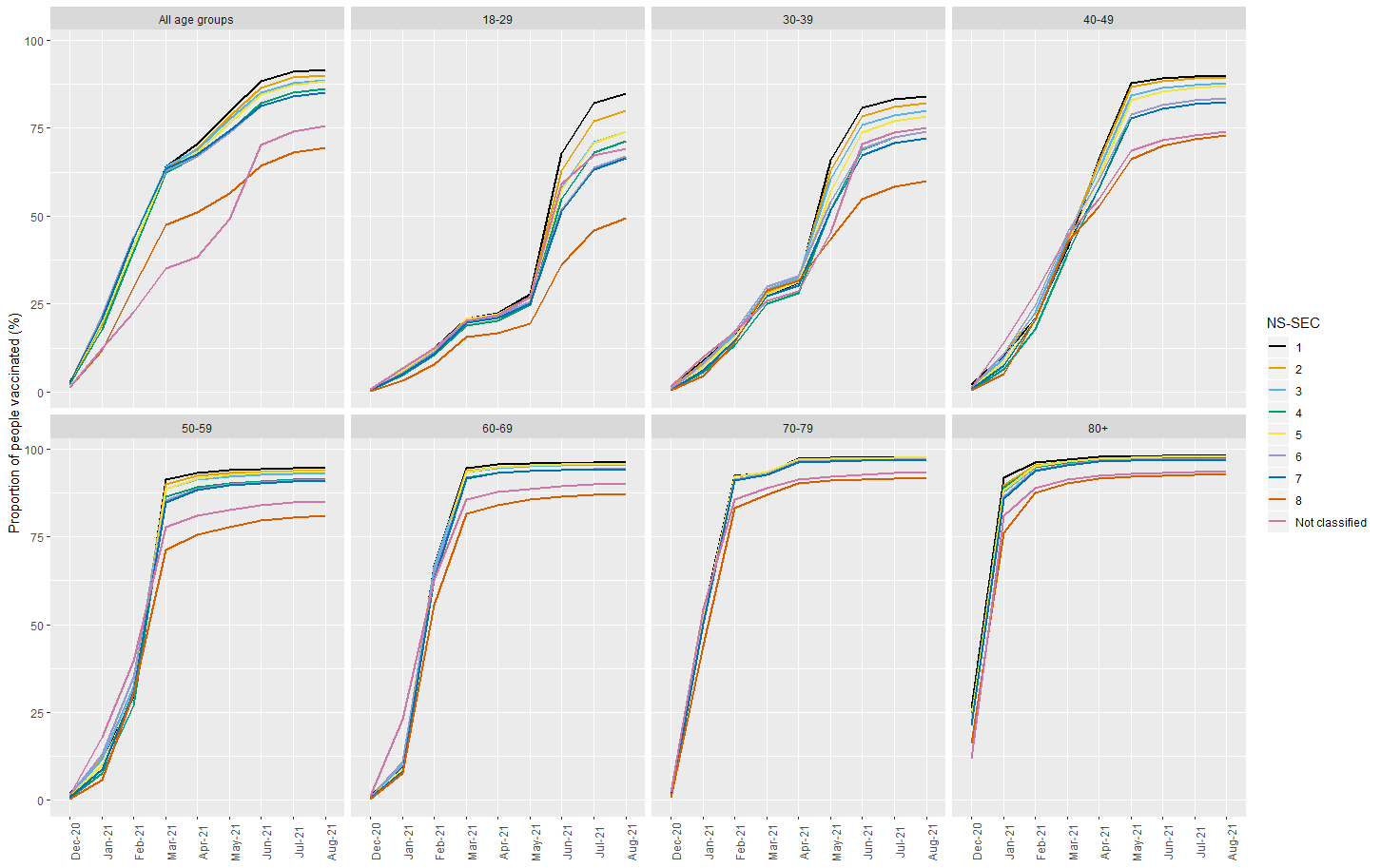

Figure S4. Proportion of people who received at least one dose of COVID-19 vaccination over time, by educational attainment and age group.

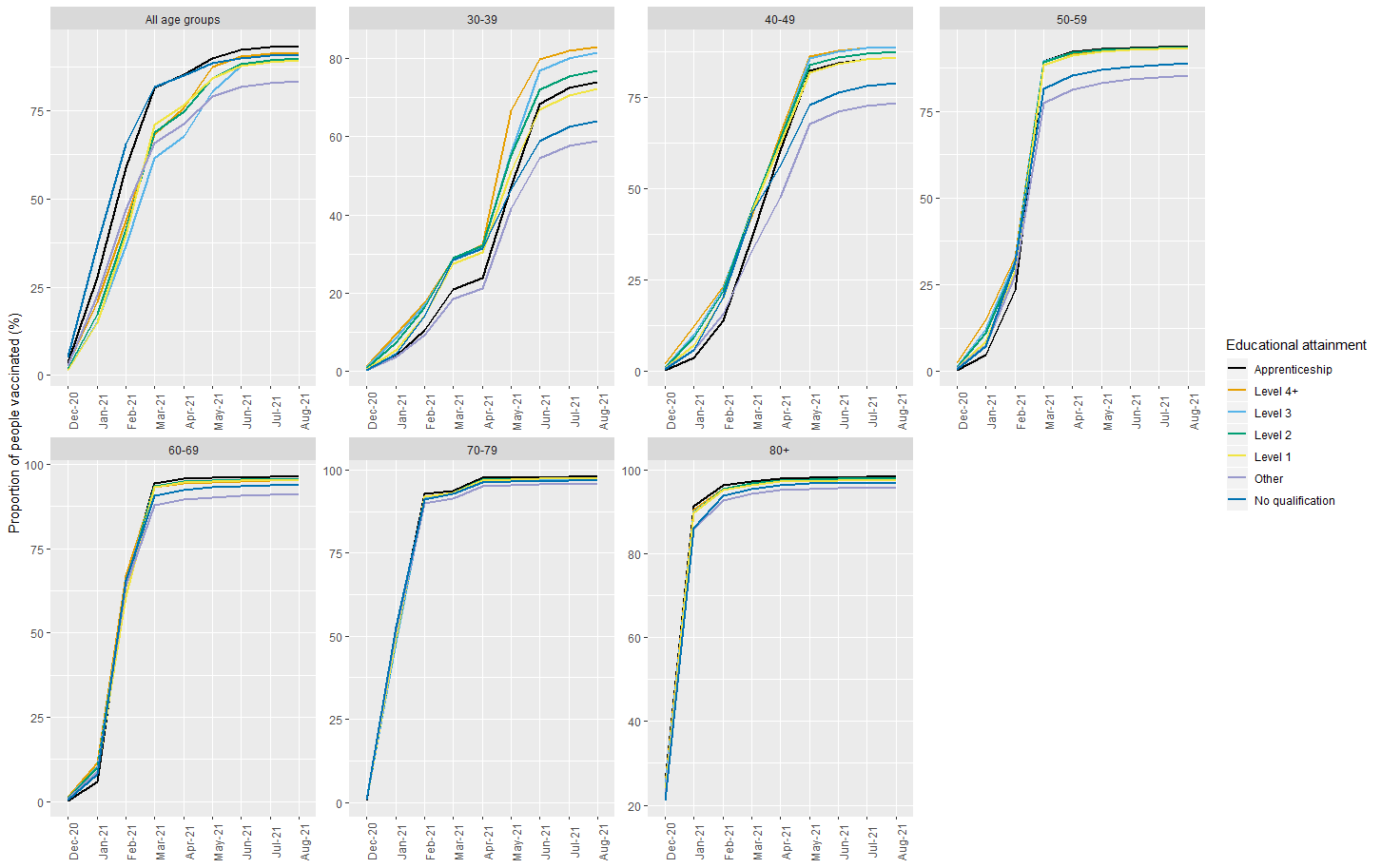

Note. Data on educational attainment are only available for those aged 30 years and over, since the 18-29 year age group were likely to have not yet completed their education in 2011 when education data were collected. Level 4+ is degree or equivalent; Level 3 A-level or equivalent, Level 2 GCSE or equivalent
